# Self-harm presentations to Emergency Departments and Place of Safety during the ‘first wave’ of the UK COVID-19 pandemic: South London and Maudsley data on service use from February to June 2020

**DOI:** 10.1101/2020.12.10.20247155

**Authors:** Eleanor Nuzum, Evangelia Martin, Gemma Morgan, Rina Dutta, Christoph Mueller, Catherine Polling, Megan Pritchard, Sumithra Velupillai, Robert Stewart

## Abstract

The lockdown and social distancing policy imposed due to the COVID-19 pandemic has had a substantial impact on both mental health service delivery, and the ways in which people are accessing these services. Previous reports from the South London and Maudsley NHS Trust (SLaM; a large mental health service provider for around 1.2m residents in South London) have highlighted increased use of virtual contacts by mental health teams, with dropping numbers of face-to-face contacts over the first wave of the pandemic. There has been concern that the impact of the COVID-19 pandemic would lead to higher mental health emergencies, particularly instances of self-harm. However, with people advised to stay at home during the ‘first wave’ lockdown, it is as yet unclear whether this impacted mental health service presentations. Taking advantage of SLaM’s Clinical Records Interactive Search (CRIS) data resource with daily updates of information from its electronic mental health records, this paper describes overall presentations to Emergency Department (ED) mental health liaison teams, and those with self-harm. The paper focussed on three periods: i) a pre-lockdown period 1^st^ February to 15^th^ March, ii) a lockdown period 16^th^ March to 10^th^ May and iii) a post-lockdown period 11^th^ May to 28^th^ June. In summary, all attendances to EDs for mental health support decreased during the lockdown period, including those with self-harm. All types of self-harm decreased during lockdown, with self-poisoning remaining the most common. Attendances to EDs for mental health support increased post-lockdown, although were only just approaching pre-lockdown levels by the end of June 2020.

## Background

The COVID-19 pandemic is suspected to have had a widespread impact on people’s mental health. For example, ‘lockdown’ and wider social distancing policies are likely to have resulted in many people being both physically and psychologically distanced from sources of support. While the main concerns have been related to increased stress and anxiety in the general population, a statement from the World Health Organisation highlighted particular concern, anticipating increased self-harm and suicidal behaviour during the pandemic (1). Within the UK, preliminary reports highlighted a shift from face-to-face to virtual contacts across community mental health services (2) and a reduction in inpatient care (3) during the first wave of the pandemic. However, less is known about the ways in which presentations to Emergency Departments (EDs) and Places of Safety (POS) were changing during this time, particularly in relation to episodes of self-harm.

We have previously been using the Clinical Records Interactive Search tool (CRIS) data platform to access anonymised electronic mental health records for individuals under the care of the South London and Maudsley NHS Foundation Trust, a large mental healthcare provider to a geographic catchment area in south London. Previous first-wave era reports are listed on https://www.maudsleybrc.nihr.ac.uk/facilities/clinical-record-interactive-search-cris/covid-19-publications/. In the present paper we used routinely extracted information to describe the pattern of adult presentations to A&E and POS for self-harm across three key periods: prior to lockdown (1^st^ February to 15^th^ March), during lockdown (16^th^ March to 10^th^ May) and after lockdown (11^th^ May to 28^th^ June). We also report on the distribution between different types of self-harm.

## Methods

### Data source

The Biomedical Research Centre (BRC) Case Register at the South London and Maudsley NHS Foundation Trust (SLaM) has been described previously (4;5). SLaM serves a geographic catchment of four south London boroughs (Croydon, Lambeth, Lewisham, Southwark; population around 1.2 million) and has used a fully electronic health record (EHR) across all its services since 2006. SLaM’s BRC Case Register was set up in 2008, providing researcher access to de-identified data from SLaM’s EHR via the Clinical Record Interactive Search (CRIS) platform and within a robust security model and governance framework (6). CRIS has been extensively developed over the last 10 years with a range of external data linkages and natural language processing resources (5) and has supported over 200 peer reviewed publications to date. Of relevance to its recent use for monitoring mental health service use during the COVID-19 pandemic, CRIS is updated from SLaM’s EHR every 24 hours and thus provides relatively ‘real-time’ data. CRIS has received approval as a data source for secondary analyses (Oxford Research Ethics Committee C, reference 18/SC/0372).

### Definition of self-harm

The methods to create a dataset of all ED presentations following self-harm were modelled on those used in previous studies using CRIS which have been previously published (7)^1^. In order to identify self-harm occurrences, we used the National Institute of Health and Social Care (8) definition: “any act of self-poisoning or self-injury carried out by an individual, irrespective of their motivation”. Any self-harm act completed within seven days of attending A&E or POS were included. Self-harm was divided into three subtypes: i) self-poisoning; ii) self-injury; iii) combinations and/or other method. Self-poisoning included any events where the individual consumed more than the recommended dose of non-recreational drugs, or where the individual consumed recreational drugs and stated that this was with the intention to self-harm. Self-injury events included any intentionally self-inflicted injuries, including cutting, stabbing, burning etc. Combination and other events included events where an individual had presented with both self-poisoning and self-injury, as well as other forms of self-harm including attempted hanging, running into traffic and jumping from a height. Attempted hanging and jumping from heights were counted as self-harm even if no actual injuries were present. Running into traffic was only counted if an actual injury was sustained.

### Case identification

CRIS was used to extract the free text entries or structured assessment forms related to any event which involved input from an A&E or POS mental health liaison team between 1^st^ February to 28^th^ June 2020 in patients aged 18 years or over. Cases of self-harm were ascertained by manual inspection of all entries which contained a keyword relating to suicidality or self-harm extracted by the CRIS query (Box 1). The free text entry that was completed for each attendance was read by at least one of three researchers (E.N., E.M. & G.M.). In order to ensure inter-rater reliability, 100 attendances were coded by all researchers, which found acceptable levels of agreement between researchers (*k*= 0.75, 0.85, 0.88 for coding relevant events, self-harm events and type of self-harm event respectively). Each attendance was coded in order to determine three criteria:

1. Relevant Event: A relevant event was defined as an event which involved an individual presenting to either A&E or Place of Safety for the attention of the mental health liaison team. Some entries extracted by CRIS related to individuals already admitted to ward beds for reasons unrelated to mental health who were being seen by a liaison team. These were not coded as relevant events and excluded from further analysis.
2. Self-Harm: Self-Harm was coded as present when the event described a self-harm incident (as defined above) that had occurred in the last seven days, where the current episode not been previously assessed in an ED or POS (to avoid double counting, for example where a patient was transferred from the POS to an ED).
3. Type: If self-harm was coded as present, the researcher then determined the type of self-harm that had occurred and coded this into the three groups previously described: self-poisoning, self-injury or combination & other.

#### Box 1. Regular expressions for self-harm and suicidality keywords in data extraction

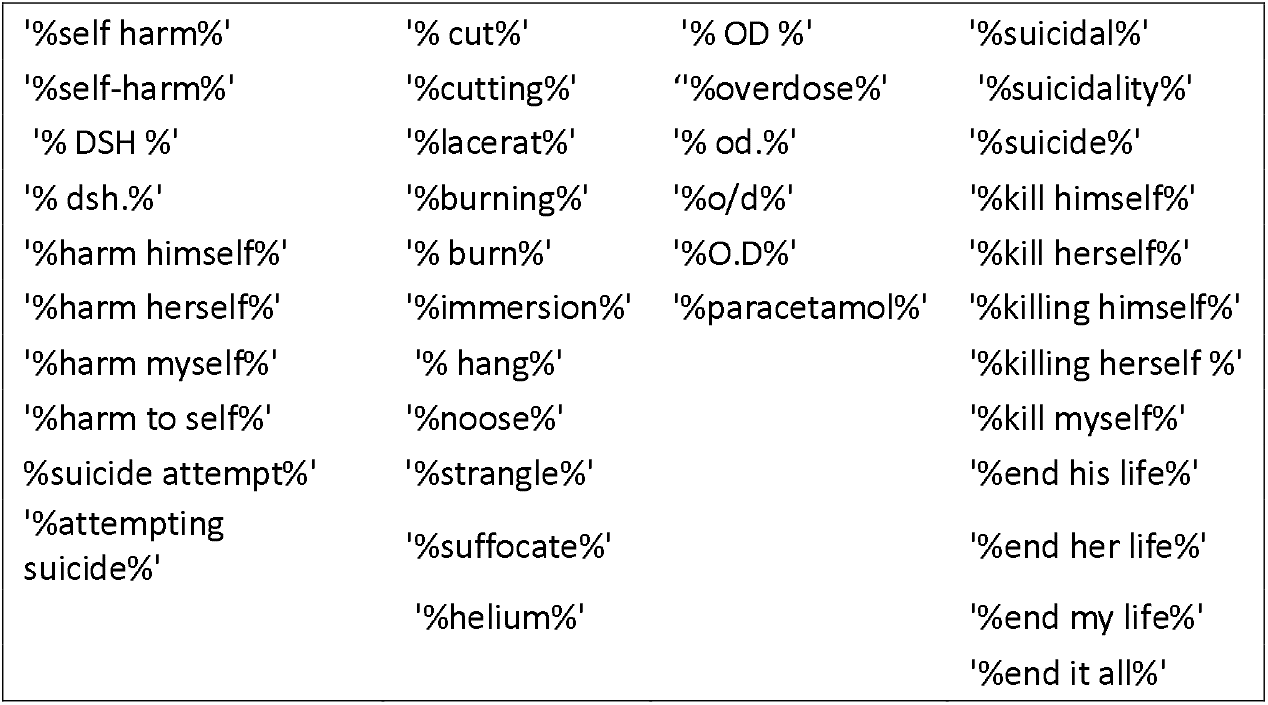

Where attendances had multiple free-text entries, the researchers only coded these once in order to ensure that no self-harm events were duplicated. In cases where individuals had more than one attendance to ED/POS for the same injury, the researchers only recorded the first attendance which reported the injury and did not count further attendances as self-harm except when it was related to a new, unreported act.

## Results

Mean weekly numbers of attendances before and during lockdown (3^rd^ Feb to 15^th^ March and 16^th^ March to 10^th^ May respectively), and proportional changes are displayed in Table 1. Mean total weekly A&E/POS events fell by 24%, and self-harm events decreased by 34%. Of self-harm events, mean weekly self-poisoning events dropped by 33%, self-injury events fell by 38% and combined/other self-harm events dropped by 36%.

**Table 1.** Weekly Presentation Numbers Before and During Lockdown (3^rd^ Feb to 15^th^ March, and 16^th^ March to 10^th^ May respectively)

| Presentations | Mean (SD)<br>before lockdown | Mean (SD) during<br>lockdown | Difference | Percentage<br>difference |
| --- | --- | --- | --- | --- |
| Total | 262 (32) | 200 (43) | -62 | -24% |
| Self-Harm | 82 (13) | 54 (14) | -28 | -34% |
| Self-Poisoning | 46 (5) | 31 (7) | -15 | -33% |
| Self-Injury | 26 (8) | 16 (6) | -10 | -38% |
| Combined/Other Self-Harm | 11 (2) | 7 (3) | -4 | -36% |

Mean differences in weekly service use during and after lockdown (16^th^ March to 10^th^ May and 11^th^ May to 28^th^ June respectively) and proportional changes are displayed in Table 2. Mean total weekly ED/POS events after lockdown were higher by 27% compared to those during lockdown, and mean weekly self-harm events were higher by 37%. More specifically, self-poisoning events had increased by 35%, self-injury events by 43% and combined/other self-harm events by 33%.

**Table 2.** Weekly□Presentation Numbers During and After Lockdown (16^th^□March to 10^th^□May and 11^th^□May□to 28^th^ June respectively)□.

| Presentations | Mean (SD)<br>during lockdown | Mean (SD) after<br>lockdown | Difference | Percentage<br>difference |
| --- | --- | --- | --- | --- |
| Total | 200 (43) | 253 (18) | 53 | +27% |
| Self-Harm | 54 (14) | 74 (9) | 20 | +37% |
| Self-Poisoning | 31 (7) | 42 (6) | 11 | +35% |
| Self-Injury | 16 (6) | 23 (6) | 7 | +43% |
| Combined/Other Self-Harm | 7 (3) | 9 (2) | 2 | +33% |

Comparisons of weekly service use before lockdown initiation and after lockdown lifting (3rd Feb to 15th March and 11th May to 28^th^ June respectively) are shown in Table 3. Mean total events were lower by 3% after lockdown, mean self-harm events were lower by 10%, self-poisoning events were lower by 9%, self-injury by 12% and combined/other self-harm events by 18%.

**Table 3.** Weekly□Presentation Numbers□Before□and After Lockdown (3^rd^□Feb to 15^th^□March and 11^th^□May□to 28^th^ June respectively)

| Presentations | Mean (SD)<br>before lockdown | Mean (SD) after<br>lockdown | Difference | Percentage<br>difference |
| --- | --- | --- | --- | --- |
| Total | 262 (32) | 253 (18) | -9 | -3% |
| Self-Harm | 82 (13) | 74 (9) | -8 | -10% |
| Self-Poisoning | 46 (5) | 42 (6) | -4 | -9% |
| Self-Injury | 26 (8) | 23 (6) | -3 | -12% |
| Combined/Other Self-Harm | 11 (2) | 9 (2) | -2 | -18% |

Further displays of the trends in weekly presentations and period differences are provided in Figures 1-4.

**Figure 1.**
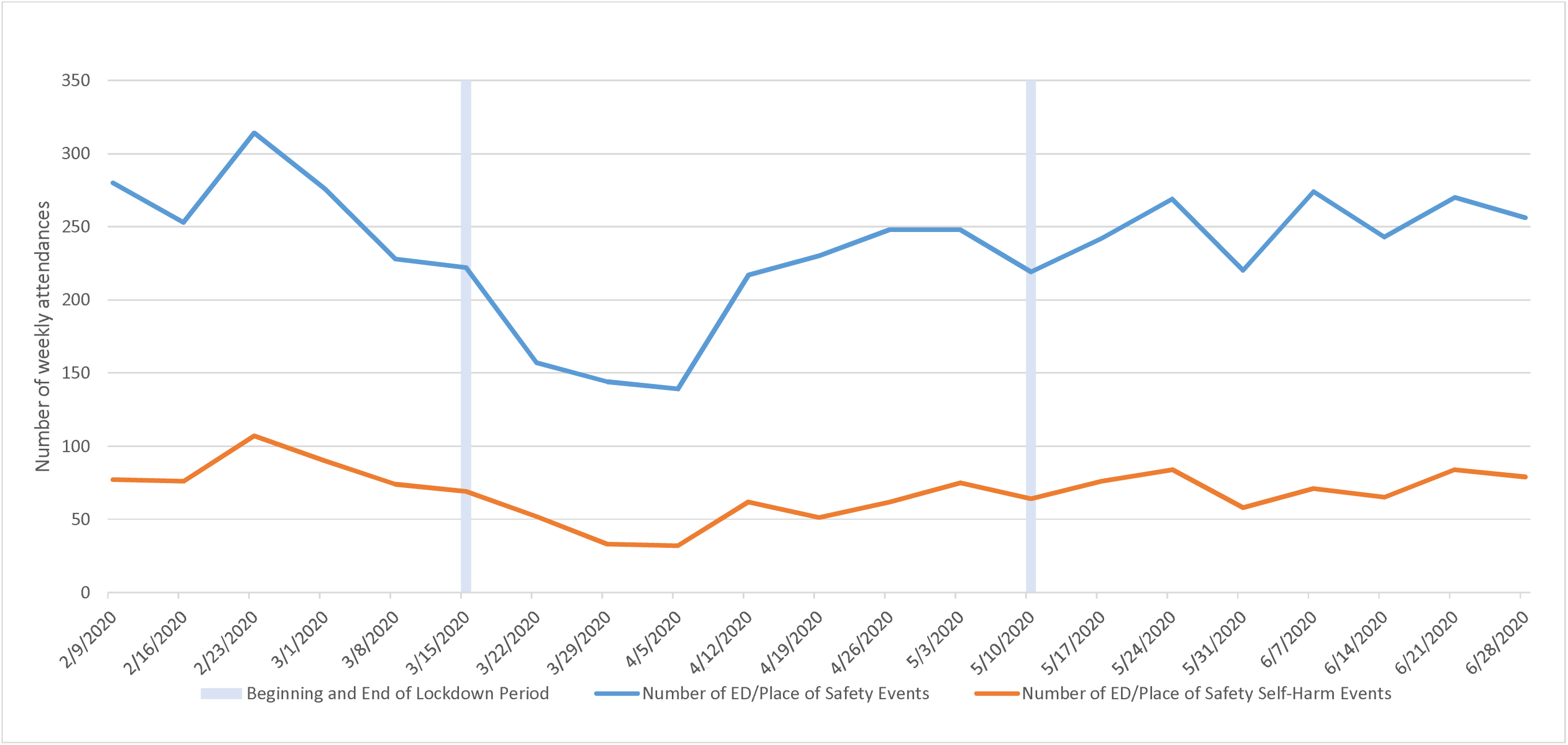
ED/Place of Safety Events (Weekly; 3rd February - 28th June 2020)

**Figure 2.**
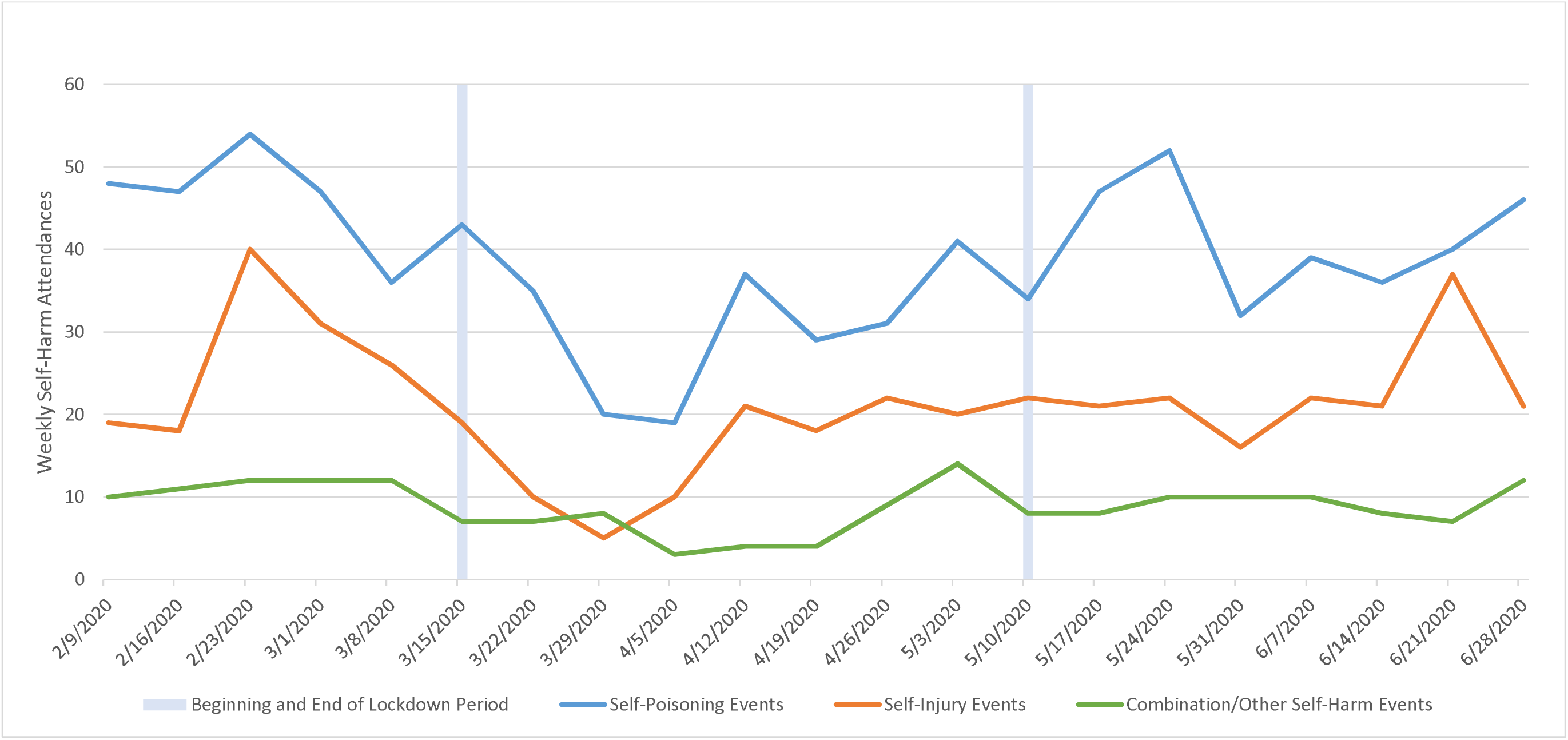
Number of Self-Harm Attendances by Type (Weekly; 3rd February - 28th June 2020)

**Figure 3.**
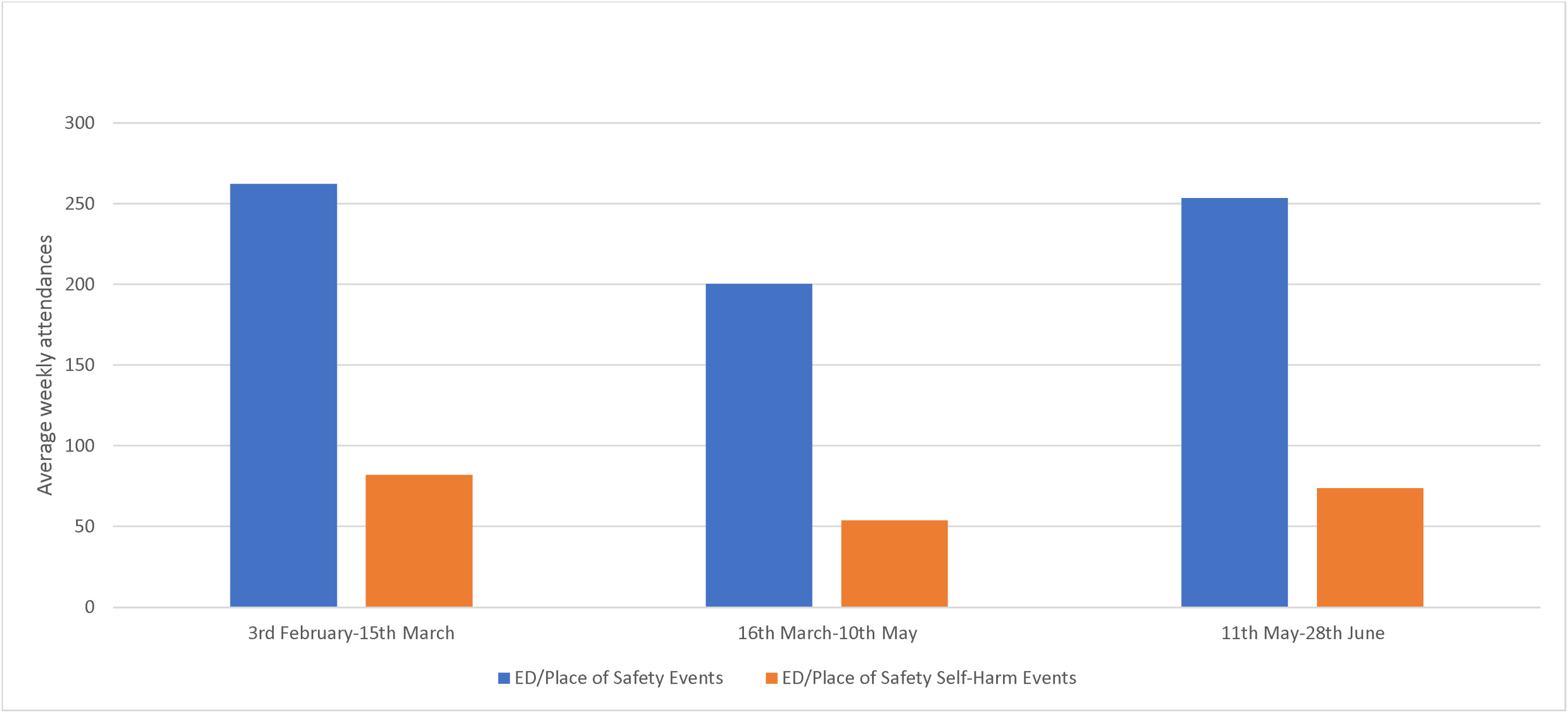
Mean Weekly ED/Place of Safety Attendances Before, During and After Lockdown Measures (3rd February - 28th June 2020)

**Figure 4.**
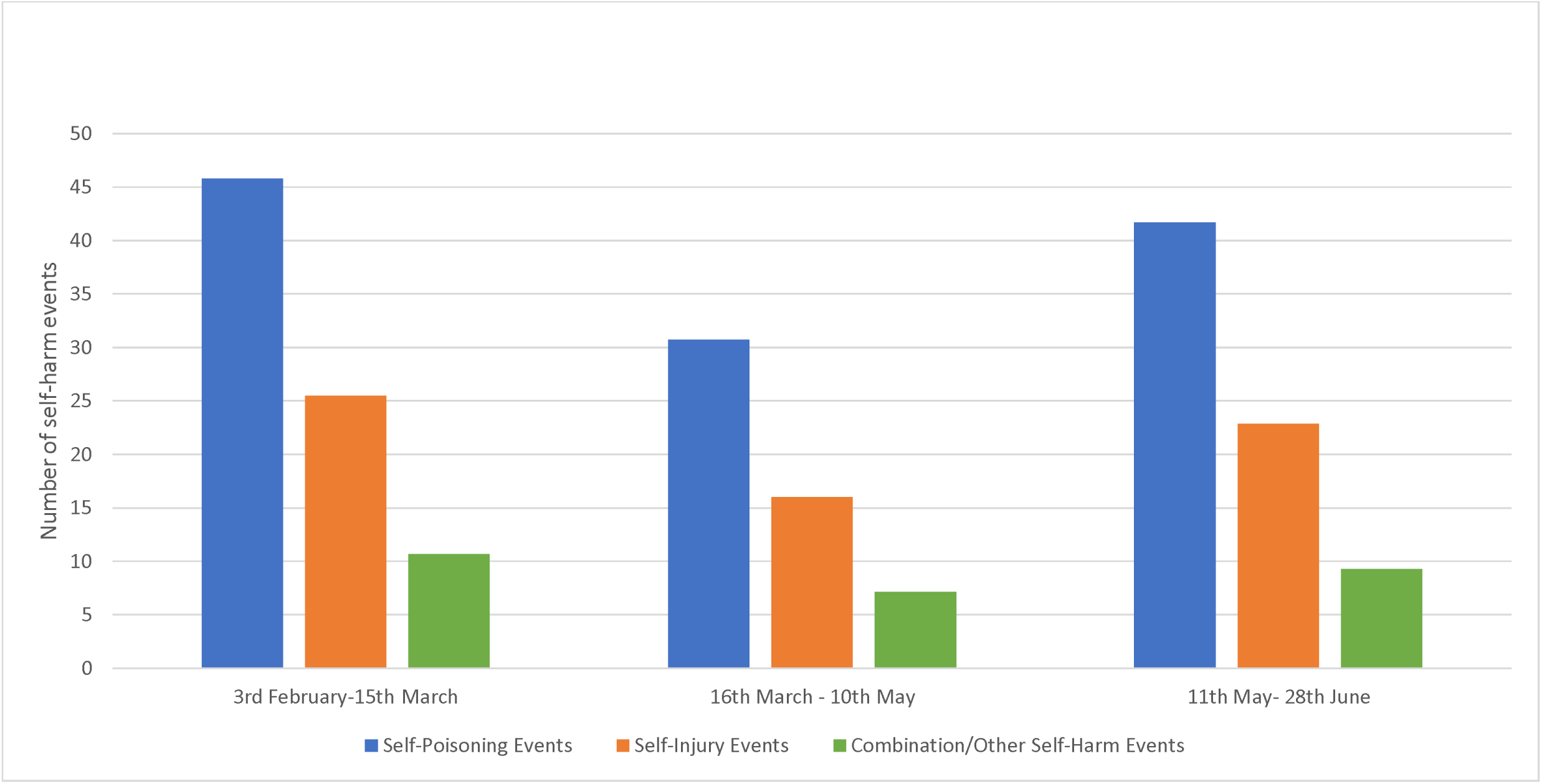
Mean Weekly Self-Harm Events by Type Before, During and After Lockdown Restrictions (3rd February - 28th June 2020)

## Discussion

The report presented here describes changes in the level of emergency mental health service use in a southeast London catchment, and self-harm presentations specifically, covering the period from February to June 2020 before and after lockdown for the ‘first wave’ of the UK COVID-19 pandemic. In summary, both presentations overall and those with self-harm were lower during lockdown, followed by an increase to levels which were not yet at pre-lockdown levels by the end of June 2020. These trends were observed for all three categories of self-harm evaluated, although were more marked for specific self-poisoning and self-injury presentations than for combination/other events.

Considering limitations, it is important to note that these data are derived from only four, neighbouring general hospitals. Because complete data are being provided for that site with no hypothetical source population intended, calculation of confidence intervals was not felt to be appropriate for the descriptive data provided in this report; applicability to other mental healthcare providers cannot therefore be inferred and would need specific investigation, although similar trends have been reported from at least one other UK site (9). It is important to remember that even outside of lockdown, the majority of self-harm in the community does not present to medical services so this data cannot be assumed to represent trends in all self-harm, only episodes resulting in emergency care (10). The services used by people for self-harm may be broader than ED and POS, which this study would not have captured and may also differ from other sites. This study also did not capture self-harm attendances to ED that did not involve ED mental health liaison team. While all four hospitals included had 24-hour liaison teams and a policy of referring and recording all attendances for self-harm throughout the period, some attendance may have been missed, either because they were not referred to mental health liaison teams at all or because they were too unwell for any mental health input throughout their time in the ED and were referred for liaison follow-up during an inpatient admission instead. Routine admissions data was not available to detect these cases, in contrast to previously published self-harm datasets produced using CRIS (7). Finally, this study only considered attenders aged 18 and over, although similar trends have been reported for children and adolescents with self-harm (10).

## Supporting information

STROBE checklist

## Data Availability

Individual patient data are required to remain within the secure network of the data provider, but can be made accessible on request from the corresponding author and subject to data provider information governance approval.

## Footnotes

1 The methods used to code CRIS data match those used in this study, but because real-time data from Hospital Episodes Statistics was not available this could not be added to the dataset in the same way it was in the published study.

